# Multiple Sclerosis Cognitive Scale (MSCS): A Brief Psychometrically-Robust Metric of Patient-Reported Cognitive Difficulty

**DOI:** 10.1101/2024.04.29.24306587

**Authors:** James F. Sumowski, Joshua Sandry

**Affiliations:** Corinne Goldsmith Dickinson Center for Multiple Sclerosis, Department of Neurology, Icahn School of Medicine at Mount Sinai; Department of Psychology, Montclair State University

## Abstract

**Background:** Cognitive decline is common in multiple sclerosis (MS), but existing metrics of patient-reported cognitive difficulties are lengthy, lack psychometric rigor, and/or fail to query expressive language deficits prevalent in MS.

**Objective:** To develop the Multiple Sclerosis Cognitive Scale (MSCS) as a brief psychometrically-robust metric of patient-reported cognitive deficits.

**Method:** Exploratory factor analysis (EFA) was conducted on 20 items of the Perceived Deficits Questionnaire (PDQ) plus five newly developed language questions in a large sample of patients with MS and matched respondents without neurologic disease. Confirmatory principal components analysis (PCA) in an independent sample assessed the EFA factor structure. Reliability of the new scale and subscales was assessed, and we evaluated the relationship between the new scale and objective cognitive impairment.

**Results:** EFA in patients (n=502) and controls (n=350), item analyses, and confirmatory PCA in an independent sample of patients (n=361) and controls (n=150) supported construction of an eight-item scale with four 2-item subscales assessing Executive / Speed, Working Memory, Expressive Language, and Episodic Memory. Internal consistency was excellent for the total MSCS (α=0.93) and good for each subscale (Executive / Speed, α=0.85; Working Memory, α=0.83, Expressive Language, α=0.87; Episodic Memory, α=0.85). There was a medium-sized relationship between objective cognitive impairment and MSCS scores (η^2^ [95%CI] = 0.06 [0.01, 0.13], but cognitive impairment was not related to the traditional PDQ (0.01 [0.00, 0.06]).

**Conclusion:** The MSCS is supported as a brief, psychometrically-robust, reliable, and valid metric of patient-reported cognitive deficits in MS. The MSCS holds promise for improving assessment of MS cognitive dysfunction in clinical and research settings.

## INTRODUCTION

Cognitive decline is common in multiple sclerosis (MS);(1) it is therefore important to have psychometrically validated, clinically-feasible patient-report tools to screen for cognitive deficits. One option is the Perceived Deficits Questionnaire (PDQ),(2) which is a 20-item scale developed in 1990 with four a priori subscales (attention/concentration, planning/organization, retrospective memory, prospective memory). The PDQ has limitations: subscales were not validated by factor analytic studies, there are no questions about expressive language deficits although word-finding difficulty is frequently reported by patients,(3) and the relatively long length of the PDQ reduces clinical feasibility due to patient burden, especially when included within a wider collection of questionnaires (e.g., fatigue, mood, physical disability). Another option is the MS Neuropsychological Questionnaire (MSNQ),(4) which is a 15-item scale developed in 2003 without cognitive subscales; items focus heavily on attention / executive function and memory without assessment of word-finding difficulty. It is also notable that the MSNQ and PDQ were developed over two and three decades ago, respectively. Others have more recently recommended Neuro-QoL short forms including a brief questionnaire assessing communication difficulties;(5) however, even these lack questions on word-finding difficulty. The current study aims to develop a brief, reliable, clinically-useful measure of patient-reported cognitive difficulties with cognitive subscales validated with factor analytic techniques.

## METHODS

### Sample

The Corinne Goldsmith Dickinson Center for Multiple Sclerosis at Mount Sinai Hospital is a tertiary care center with a catchment area encompassing the racially/ethnically diverse New York Metropolitan Area. In 2018, we established a clinic aiming to perform cognitive screenings for all patients at our center; the 20-item PDQ plus five language questions was completed by patients from August 2018 through August 2021. We performed an IRB approved retrospective chart review of clinical and patient-reported cognitive data from all patients aged 18 to 65 years and diagnosed with relapse-onset MS from 1995 onward who completed cognitive screenings. Self-reported cognitive difficulty was also collected from demographically-matched persons without neurologic conditions through an IRB-exempt anonymous electronic capture questionnaire.

### PDQ plus Language Questions

The PDQ is a 20-item self-report inventory assessing frequency of cognitive difficulties as never (0), rarely (1), sometimes (2), fairly often (3), or very often (4). The PDQ has four *a priori* subscales (with five items each) assessing attention/concentration, planning/organization, retrospective memory, and prospective memory, but there is no assessment of language difficulty. Our development of five language questions to add to the PDQ was informed by clinical experience with patients and confirmed by retrospective chart review of self-report remote electronic capture (REDCap) questionnaire in consecutive patients (meeting aforementioned inclusion criteria) during a 12-month period. Patients were asked “Do you have any concerns about your current cognition?” If yes, an open field question displayed: “In a few words, please describe the cognitive problem(s) that you experience.” These questions were completed prior to completion of the PDQ to avoid biasing results. Three independent raters (see acknowledgements) reviewed all free text responses to identify presence or absence of self-reported language difficulty, and then to categorize difficulties into like categories.

### Statistical Approach

Exploratory factor analysis was performed on responses to the 25 questions by patients and controls using R version 4.2.1 and the psych package.(6) The goal was to develop an empirically-derived brief cognitive screener; as such, we identified the two items with the highest factor loadings and highest internal consistency (Cronbach’s alpha) for each factor, which were selected for the new brief screener. Next, confirmatory principal components analysis was performed for selected items in an independent sample of patients and controls.

## RESULTS

### Sample

Data on the 25-item self-report cognitive questionnaire were captured from 502 persons with MS and 350 demographically-matched control respondents for the exploratory factor analysis (Table 1).

**Table 1.** Sample Characteristics.

|  | **EFA Sample (n=852)** |  | **Confirmatory PCA Samples (n=511)** |  |  |
| --- | --- | --- | --- | --- | --- |
|  | **MS**<br>**(n=502)** | **Controls**<br>**(n=350)** | **Research**<br>**MS (n=167)** | **Clinical MS**<br>**(n=194)** | **Controls**<br>**(n=150)** |
| Age (years), mean [SD] | 43.6 [11.5] | 44.3 [12.5] | 36.2 [7.5] | 42.9 [10.8] | 43.5 [14.0] |
| Sex, N [%] |  |  |  |  |  |
| Women | 357 [71.1] | 252 [72.0] | 111 [66.5] | 147 [75.8] | 108 [72.0] |
| Men | 145 [28.9] | 98 [28.0] | 56 [33.5] | 47 [24.2] | 42 [28.0] |
| Bachelor's degree, N [%] | 373 [74.3] | 275 [78.6] | 130 [77.8] | 128 [66.0] | 85 [56.7] |
| MHI-5, mean [SD] | 70.5 [15.8] | 76.5 [13.0] | 72.4 [15.8] | 69.9 [15.1] | 73.5 [13.2] |
| Race and Ethnicity, N [%] |  |  |  |  |  |
| Black, (not Hispanic or Latino/a) | 82 [16.3] |  | 24 [14.4] | 53 [27.3] |  |
| Hispanic or Latino/a | 87 [17.3] |  | 38 [22.8] | 45 [23.2] |  |
| White (not Hispanic or Latino/a) | 318 [63.3] |  | 98 [58.7] | 92 [47.4] |  |
| Other (not Hispanic or Latino/a) | 15 [3.0] |  | 7 [4.2] | 4 [2.1] |  |
| Years since MS dx, median [IQR] | 7 [2, 13] |  | 4 [3, 6] | 5.5 [1, 12] |  |
| Disease Course, N [%] |  |  |  |  |  |
| RRMS | 419 [83.5] |  | 167 [100] | 175 [90.2] |  |
| SPMS | 83 [16.5] |  | 0 [0] | 19 [9.8] |  |
| Disease Modifying Therapy, N [%] |  |  |  |  |  |
| None | 73 [14.5] |  | 13 [7.8] | 23 [11.9] |  |
| Platform injectables, teriflunomide | 97 [19.3] |  | 24 [14.4] | 30 [15.5] |  |
| S1p modulators, fumarates | 142 [28.3] |  | 62 [37.1] | 48 [24.7] |  |
| Monoclonal antibodies | 190 [37.8] |  | 68 [40.7] | 93 [47.9] |  |

### Language Questions

Data were captured from a subset of 319 consecutive patients with MS for validation of language questions. Of 319 patients, about half (51%, n=163) endorsed “yes” to having concerns about cognition; free text responses were reviewed by three independent raters to identify presence or absence of self-reported language difficulty (interrater agreement was excellent, Kappa [95%CI] of 0.85 [0.76, 0.93]). Language deficits were considered present if all three raters (n=54) or two of three raters (n=10) coded it present; language difficulty was reported by 39% of patients endorsing concerns about cognition (n=64 of 163), and 20% of all patients regardless of endorsement of cognitive concerns (n=64 of 319). Analysis of the 64 responses identified (a) word-finding difficulty (n=46) with descriptions of trouble retrieving known words (i.e., “tip of the tongue” phenomenon), (b) difficulty clearly expressing thoughts (n=13), (c) using the wrong word or misspeaking (n=10), (d) difficulties comprehending text or discourse (n=6), and (e) other and/or vague responses (n=6; e.g., “speech problems,” misspelling). Analyses support use of the five language questions (last five items in Table 2).

**Table 2.** Results of Exploratory Factor Analysis of 25 Self-Reported Cognitive Deficits. The 25 items are from the 20-item PDQ plus five new language items (in italics below).

|  | **Factors** |  |  |  |  |
| --- | --- | --- | --- | --- | --- |
| **Perceived Deficits Questionnaire Item plus Language Questions** | **A** | **B** | **C** | **D** | **E** |
| losing your train of thought | 0.13 | 0.01 | 0.29 | 0.18 | **0.46** |
| trouble remembering people's names, even familiar people | -0.09 | 0.25 | 0.28 | 0.11 | **0.35** |
| forgetting what you came into the room for | 0.07 | 0.17 | 0.30 | 0.06 | **0.47** |
| trouble getting things organized | **0.62** | 0.10 | -0.03 | -0.01 | 0.28 |
| trouble concentrating on what people are saying, or on what you are reading or watching on TV | **0.41** | -0.05 | 0.05 | 0.41 | 0.25 |
| forgetting if you had already done something | 0.15 | 0.30 | 0.15 | 0.13 | **0.33** |
| forgetting appointments or meetings you had planned | 0.16 | **0.58** | 0.00 | 0.04 | 0.22 |
| difficulty planning what to do in the day | **0.66** | 0.23 | -0.05 | 0.03 | 0.03 |
| difficulty doing more than one thing at a time | **0.54** | 0.23 | 0.07 | 0.06 | 0.01 |
| trouble remembering where you put something, like your keys or where you parked your car | 0.14 | **0.54** | 0.06 | -0.06 | 0.25 |
| forgetting what day of the week it is | 0.10 | **0.59** | 0.05 | 0.05 | -0.02 |
| trouble getting started, even if you had lots to do | **0.84** | -0.01 | 0.06 | -0.03 | -0.03 |
| taking a long time to finish things | **0.81** | 0.01 | 0.11 | -0.03 | -0.01 |
| forgetting details of a recent conversation | 0.13 | 0.18 | 0.23 | **0.37** | 0.15 |
| forgetting to do things like turning on your alarm clock, or charging your smartphone or computer | 0.04 | **0.79** | 0.03 | -0.04 | -0.06 |
| finding your mind drifting, or your mind going blank | **0.37** | 0.09 | 0.11 | 0.32 | 0.09 |
| trouble holding a string of numbers in your head, even for a few seconds | 0.04 | **0.34** | 0.09 | 0.31 | 0.18 |
| trouble recalling what happened during the last week | -0.07 | 0.43 | 0.14 | **0.44** | 0.07 |
| forgetting to do routine things without reminders, like taking your medication, or picking someone up | 0.06 | **0.72** | 0.07 | 0.05 | -0.07 |
| trouble making decisions | **0.58** | 0.10 | 0.05 | 0.23 | -0.17 |
| *having a word "on the tip of your tongue" but with difficulty getting it out* | 0.03 | 0.00 | **0.81** | -0.05 | 0.14 |
| *having to read something several times to understand it* | 0.26 | -0.02 | **0.35** | 0.32 | 0.04 |
| *having a sense of what you want to say, but you have trouble clearly expressing your thoughts* | 0.07 | 0.00 | **0.78** | 0.04 | 0.01 |
| *accidentally saying the wrong word / misspeaking* | -0.01 | 0.07 | **0.80** | -0.01 | -0.09 |
| *having difficulty following a conversation, especially a conversation with multiple people or parts* | 0.15 | 0.15 | **0.39** | 0.36 | -0.16 |

### Exploratory Factor Analysis (EFA)

Responses to the 25 questions by 852 consecutive respondents (502 patients, 350 controls) were analyzed with EFA using R version 4.2.1 and the psych package. The Kaiser-Meyer-Olkin measure of sampling adequacy was .98 suggesting excellent factorability. Results from the parallel analysis, in concordance with the scree plot, suggested that a five-factor solution with oblimin rotation has excellent fit (RMSEA = .05, TLI = .962; Table 2, Figure 1). Of the four items with highest loadings for each factor, we identified the two with the highest internal consistency (Cronbach’s alpha). For factor A, internal consistency was good for “get started when lots to do” and “take long time to finish things” (α=0.88). For factor B, internal consistency was acceptable for “forget to take medication, etc.” and “forget meetings, appointments” (α=0.79). For factor C, internal consistency was good for “word on ‘tip of tongue’” and “clearly expressing thoughts” (α=0.85). Factor D consisted of two items with good internal consistency (α=0.85). For factor E, internal consistency was good for “forget why entered room” and “losing train of thought” (α=0.85). Internal consistency was good for all two-item pairs across factors except factor B. Further inspection revealed possible floor effects for all four items with highest loadings for factor B, with < 10% of all respondents endorsing “fairly often” or “very often.” These items are four of the five items of the PDQ prospective memory subscale; means for this scale were much lower than all other PDQ subscales in the original publication.(2) Only one of the other 21 questions showed a possible floor effect (“follow complex conversations”). To derive a brief scale with the best reliability and clinical relevance, we excluded factor B. EFA with the remaining eight items yielded four factors (Table 3, Figure 2), which are best characterized as Executive / Speed, Episodic Memory, Working Memory, and Expressive Language.

**Figure 1.**
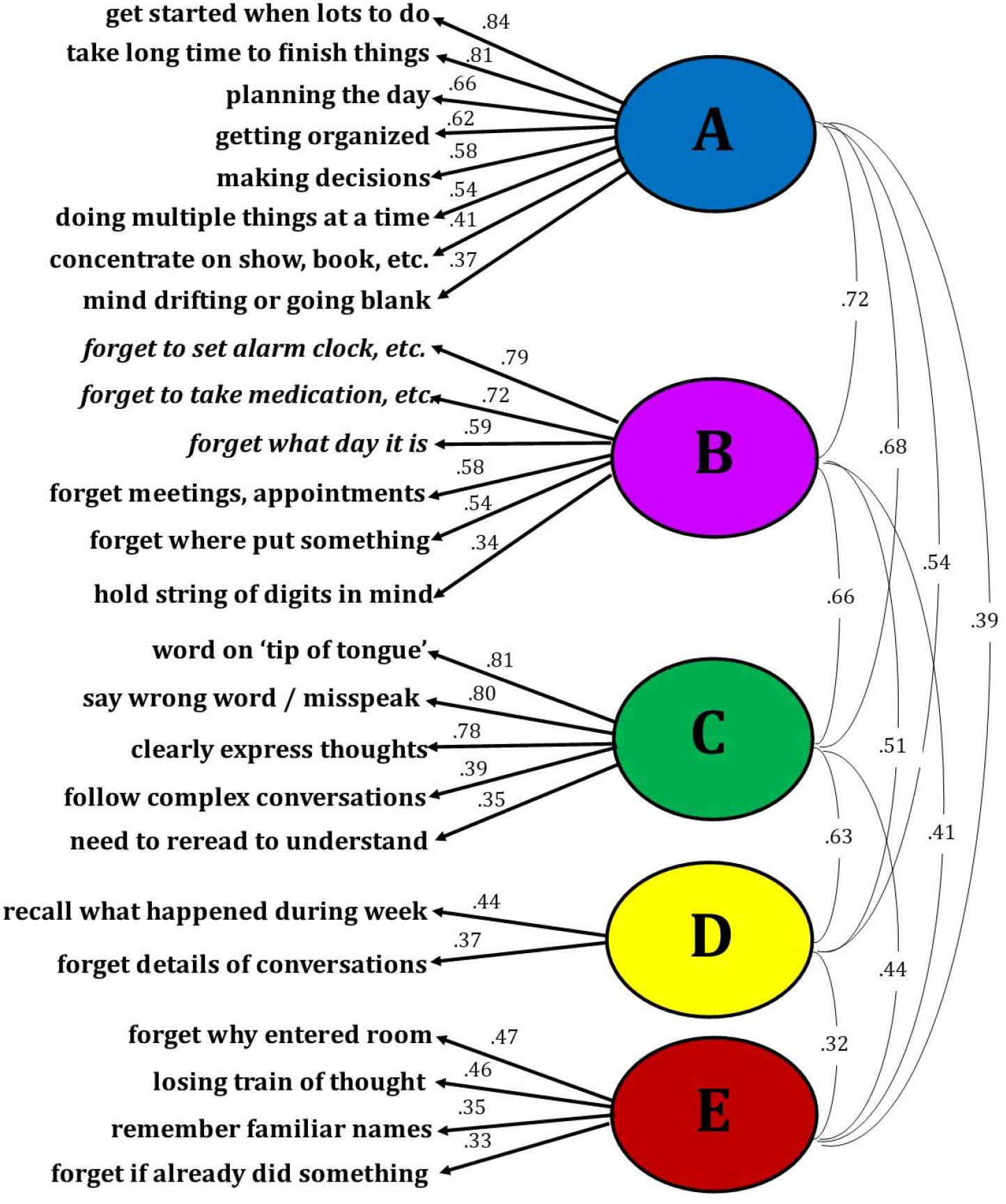
Exploratory Factor Analysis of 25 Self-Reported Cognitive Deficits.

**Table 3.** Results of Exploratory Factor Analysis of Eight MS Cognitive Scale (MSCS) Items.

|  | **Current Sample w/ PDQ (n=852; 502 MS, 350 HC)** |  |  |  |
| --- | --- | --- | --- | --- |
| **MSCS Item** | **Executive /**<br>**Speed (ES)** | **Working**<br>**Memory (WM)** | **Expressive**<br>**Language (EL)** | **Episodic**<br>**Memory (EM)** |
| trouble getting started | **0.88** | 0.04 | -0.02 | -0.04 |
| take a long time to finish things | **0.88** | -0.04 | 0.03 | 0.05 |
| losing your train of thought | 0.01 | **0.79** | 0.02 | 0.05 |
| forgetting why you entered a room | 0.01 | **0.84** | 0.01 | 0.00 |
| word on the tip of your tongue | 0.00 | 0.13 | **0.79** | -0.05 |
| trouble clearly expressing your thoughts | 0.02 | -0.08 | **0.85** | 0.06 |
| forgetting details of conversations | 0.04 | 0.02 | 0.00 | **0.84** |
| trouble recalling what happened during last week | -0.03 | 0.01 | 0.01 | **0.83** |

**Figure 2.**
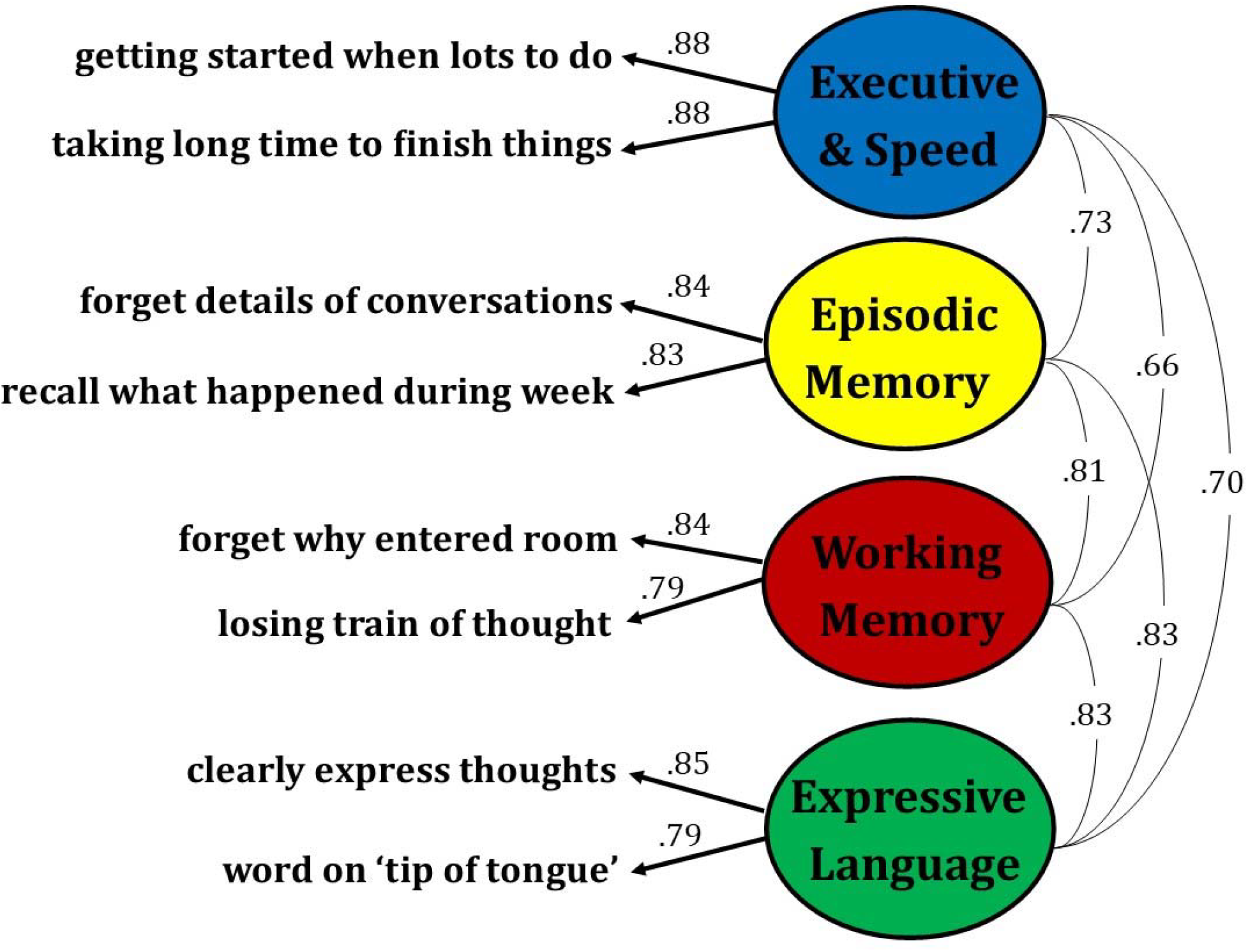
Exploratory Factor Analysis of the Eight MSCS Items.

### Confirmatory Principal Components Analyses (PCA)

Confirmatory PCAs (four components, oblimin rotation) with the eight selected items were performed in an independent sample combining a research sample of 167 persons with relapsing-remitting MS who completed the full 25-item survey, a clinical sample of 194 patients with MS who completed the brief scale, and a sample of 150 respondents without neurologic conditions who completed the brief scale. As shown (Table 4), confirmatory PCA in this independent sample supported the factor structure of the eight-item MSCS, which was also shown when performing separate PCAs for all patients (n=863) and all controls (n=500; Table 5).

**Table 4.** Results of Confirmatory Principal Component Analyses in Independent Sample.

|  | **Confirmatory PCA Sample (n=511; 361 MS, 150 HC)** |  |  |  |
| --- | --- | --- | --- | --- |
| **MSCS Item** | **Executive /**<br>**Speed (ES)** | **Working**<br>**Memory (WM)** | **Expressive**<br>**Language (EL)** | **Episodic**<br>**Memory (EM)** |
| trouble getting started | **0.93** | 0.07 | -0.02 | 0.04 |
| take a long time to finish things | **0.86** | -0.03 | 0.08 | -0.06 |
| losing your train of thought | 0.06 | **0.95** | -0.06 | -0.01 |
| forgetting why you entered a room | -0.02 | **0.74** | 0.18 | -0.08 |
| word on the tip of your tongue | 0.00 | 0.12 | **0.89** | 0.05 |
| trouble clearly expressing your thoughts | 0.15 | -0.05 | **0.78** | -0.15 |
| forgetting details of conversations | -0.07 | 0.07 | 0.16 | **-0.83** |
| trouble recalling what happened during last week | 0.08 | 0.01 | -0.09 | **-0.94** |

**Table 5.**
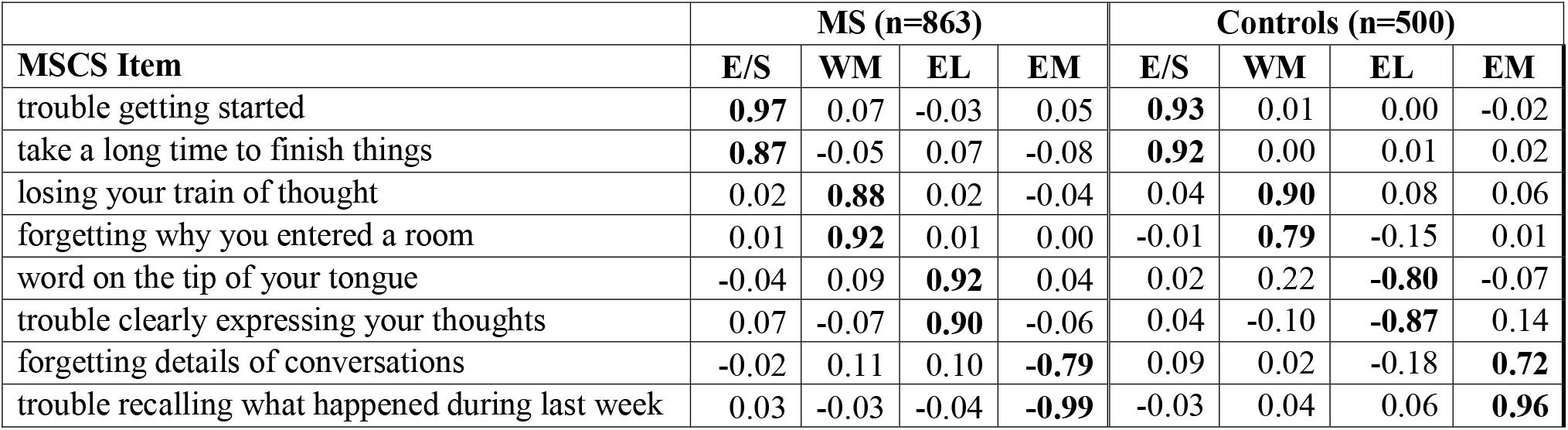
Results of Confirmatory Principal Component Analyses Separately for All Patients and All Controls.

|  | **MS (n=863)** |  |  |  | **Controls (n=500)** |  |  |  |
| --- | --- | --- | --- | --- | --- | --- | --- | --- |
| **MSCS Item** | **E/S** | **WM** | **EL** | **EM** | **E/S** | **WM** | **EL** | **EM** |
| trouble getting started | **0.97** | 0.07 | -0.03 | 0.05 | **0.93** | 0.01 | 0.00 | -0.02 |
| take a long time to finish things | **0.87** | -0.05 | 0.07 | -0.08 | **0.92** | 0.00 | 0.01 | 0.02 |
| losing your train of thought | 0.02 | **0.88** | 0.02 | -0.04 | 0.04 | **0.90** | 0.08 | 0.06 |
| forgetting why you entered a room | 0.01 | **0.92** | 0.01 | 0.00 | -0.01 | **0.79** | -0.15 | 0.01 |
| word on the tip of your tongue | -0.04 | 0.09 | **0.92** | 0.04 | 0.02 | 0.22 | **-0.80** | -0.07 |
| trouble clearly expressing your thoughts | 0.07 | -0.07 | **0.90** | -0.06 | 0.04 | -0.10 | **-0.87** | 0.14 |
| forgetting details of conversations | -0.02 | 0.11 | 0.10 | **-0.79** | 0.09 | 0.02 | -0.18 | **0.72** |
| trouble recalling what happened during last week | 0.03 | -0.03 | -0.04 | **-0.99** | -0.03 | 0.04 | 0.06 | **0.96** |

### Reliability

Internal consistency (Cronbach’s alpha) of the eight-item scale among patients who completed the brief form (n=194) was excellent for the total MSCS (α=0.93) and good for each subscale (Executive / Speed, α=0.85; Episodic Memory, α=0.85; Working Memory, α=0.83, Expressive Language, α=0.87). Test-retest reliability was assessed with intraclass correlation coefficients (ICC; two-way mixed ANOVA; absolute agreement(7)) for the brief form completed twice by 62 patients with MS, with a one-year interval between assessments; reliability was good for the total MSCS (ICC [95% CI], 0.82 [0.72, 0.89]) and for each subscale (ICCs 0.74 to 0.81; Table 6). This is likely an underestimate of the actual test-retest reliability given the long one-year interval between assessments.

**Table 6.** Test-Retest Reliabilities of MSCS Subscales in Patients completing the brief form at two time points (n=62) ICC [95%CI] with two-way mixed analysis of variance model with interaction for the absolute agreement between single scores. ICC for the total MSCS score was 0.82 [0.72, 0.89].

|  |  | TIME 1 |  |  |  |
| --- | --- | --- | --- | --- | --- |
|  |  | E/S | WM | EL | EM |
| TIME 2 | E/S | **0.81**<br>[0.70, 0.88] | 0.55<br>[0.34, 0.70] | 0.59<br>[0.38, 0.74] | 0.67<br>[0.50, 0.78] |
|  | WM | 0.58<br>[0.40, 0.73] | **0.74**<br>[0.60, 0.83] | 0.64<br>[0.47, 0.77] | 0.66<br>[0.48, 0.78] |
|  | EL | 0.67<br>[0.51, 0.79] | 0.62<br>[0.45, 0.75] | **0.74**<br>[0.59, 0.83] | 0.72<br>[0.57, 0.82] |
|  | EM | 0.68<br>[0.53, 0.80] | 0.55<br>[0.32, 0.70] | 0.54<br>[0.31, 0.70] | **0.80**<br>[0.69, 0.87] |

### Link to Objective Cognitive Performance

A brief screening battery adopted for MS (BICAMS)(8) consists of a high-sensitivity information processing task (Symbol Digit Modalities Test, SDMT)(9) and measures of word-list learning and object-location memory. Our modified version of this battery includes SDMT, word-list total learning on the Hopkins Verbal Learning Test, Revised (HVLT-R),(10) and object-location memory on CANTAB Paired Associate Learning (PAL).(11) Task performance data were available for 502 patients who completed the full 20-item PDQ and 194 patients who completed the MSCS. Raw scores were converted to age-adjusted norm-referenced z-scores relative to each test’s healthy standardization sample, which was then used to characterize impairment for each task as performance ≤1.5 standard deviations below normal (z-score ≤ -1.5). Patients were categorized as having impairment on 0, 1, or 2+ tests. We then matched patients from the larger PDQ sample to the smaller MSCS sample for age, sex, race/ethnicity, education, MS phenotype, time since diagnosis, and mood (MHI-5), resulting in extremely well-matched samples of (a) 194 patients who completed the full 20-item PDQ and (b) 194 patients who completed the MSCS (Table 7). One-way ANOVAs tested differences in the PDQ (mean of 20 items) and the MSCS (mean of eight items) across patients with impairment on 0, 1, or 2+ tests. As shown (Figure 3), MSCS differed across levels of cognitive impairment (F[2, 193]=5.85, p=0.003; η^2^ [95%CI] = 0.06 [0.01, 0.13]) whereby patient-reported difficulty was worse among patients with 2+ impaired tests than those with ≤1 impaired test. In contrast, there was no difference in PDQ across levels of cognitive impairment (F[2, 193]=1.34, p=0.265; η^2^ [95%CI] = 0.01 [0.00, 0.06]). One-way ANOVAs were repeated after adjusting MSCS and PDQ for mood (MHI-5, using GLM). Again, as shown (Figure 3), there were differences in MSCS across levels of cognitive impairment F[2, 193]= 3.83, p=0.023; η^2^ [95%CI] = 0.04 [0.00, 0.10]), but PDQ did not differ across levels of impairment (F[2, 193]=0.15, p=0.864; η^2^ [95%CI] = 0.00 [0.00, 0.02]).

**Table 7.**
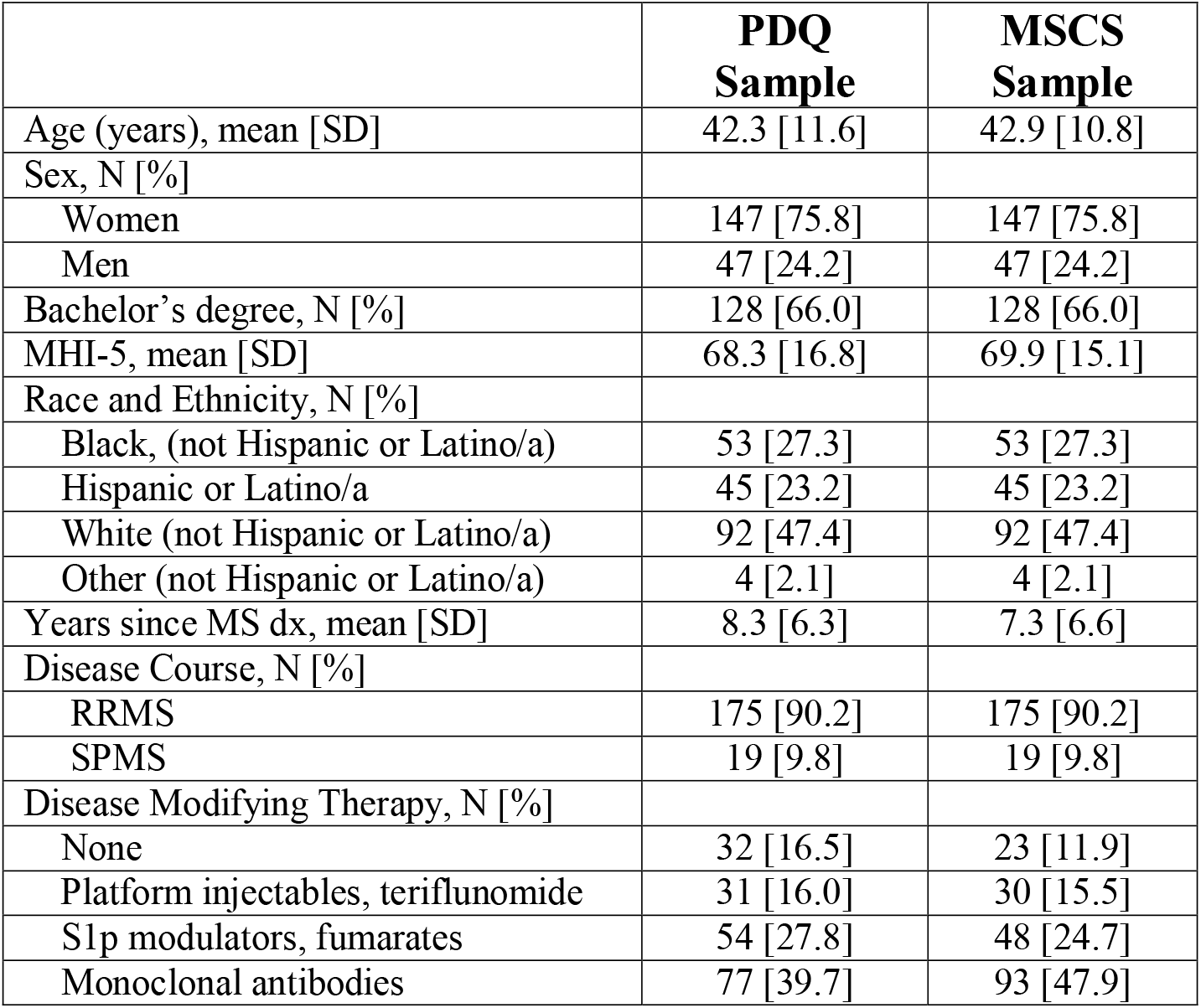
Matched samples of patients with MS who completed the PDQ or the MSCS.

|  | **PDQ**<br>**Sample** | **MSCS**<br>**Sample** |
| --- | --- | --- |
| Age (years), mean [SD] | 42.3 [11.6] | 42.9 [10.8] |
| Sex, N [%] |  |  |
| Women | 147 [75.8] | 147 [75.8] |
| Men | 47 [24.2] | 47 [24.2] |
| Bachelor's degree, N [%] | 128 [66.0] | 128 [66.0] |
| MHI-5, mean [SD] | 68.3 [16.8] | 69.9 [15.1] |
| Race and Ethnicity, N [%] |  |  |
| Black, (not Hispanic or Latino/a) | 53 [27.3] | 53 [27.3] |
| Hispanic or Latino/a | 45 [23.2] | 45 [23.2] |
| White (not Hispanic or Latino/a) | 92 [47.4] | 92 [47.4] |
| Other (not Hispanic or Latino/a) | 4 [2.1] | 4 [2.1] |
| Years since MS dx, mean [SD] | 8.3 [6.3] | 7.3 [6.6] |
| Disease Course, N [%] |  |  |
| RRMS | 175 [90.2] | 175 [90.2] |
| SPMS | 19 [9.8] | 19 [9.8] |
| Disease Modifying Therapy, N [%] |  |  |
| None | 32 [16.5] | 23 [11.9] |
| Platform injectables, teriflunomide | 31 [16.0] | 30 [15.5] |
| S1p modulators, fumarates | 54 [27.8] | 48 [24.7] |
| Monoclonal antibodies | 77 [39.7] | 93 [47.9] |

**Figure 3.**
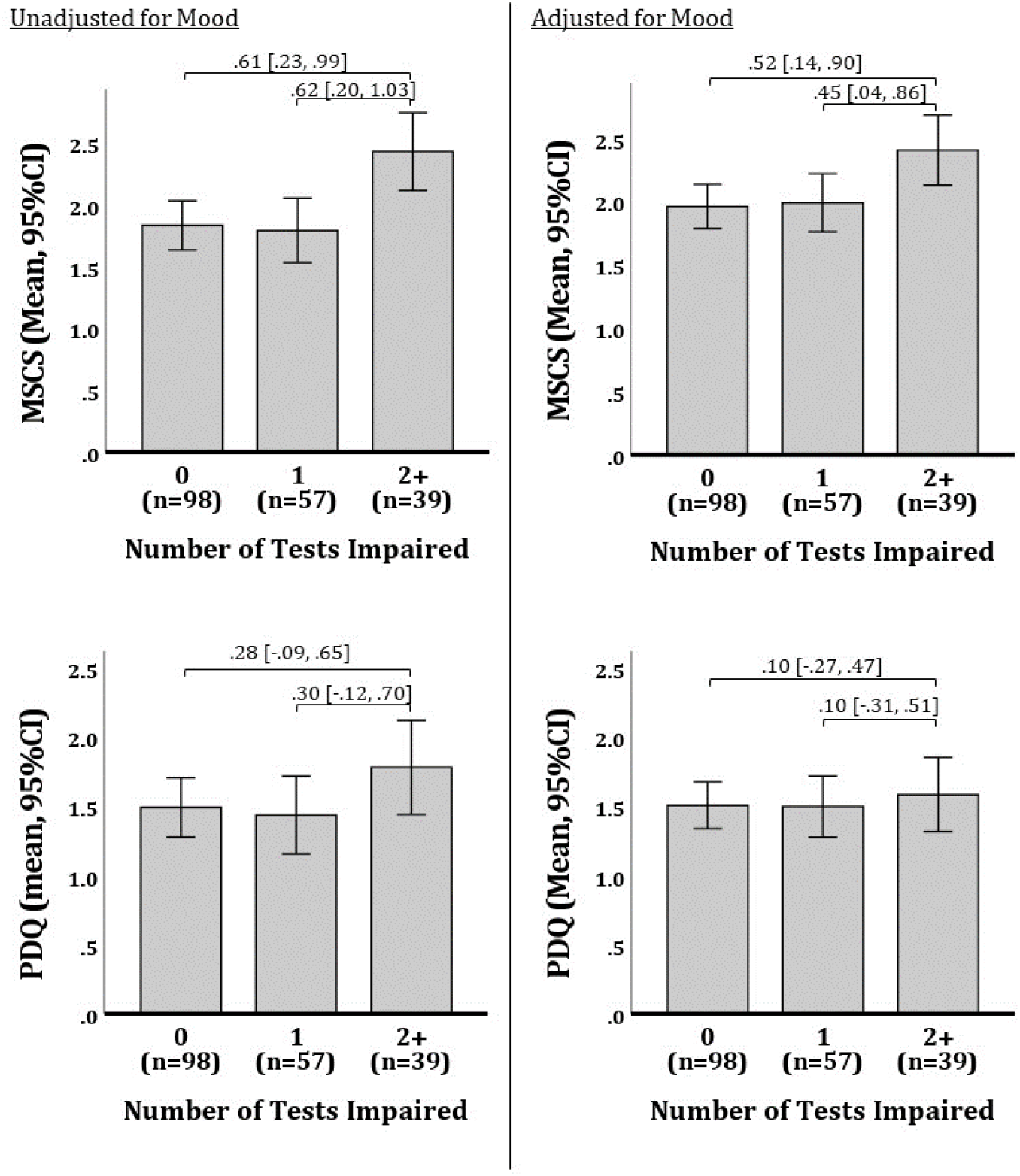
Differences in Patient-Reported Cognitive Difficulty across levels of Cognitive Impairment in Matched Samples completing the PDQ or MSCS. Results are presented for MSCS and PDQ scores before (left) and after (right) adjusting scores for mood (MHI-5). Effect sizes (Cohen’s D [95%CI]) are presented for comparisons across levels of cognitive impairment.

## DISCUSSION

We report good reliability and validity for the Multiple Sclerosis Cognitive Scale (MSCS; see Appendix), a new eight-item patient-report cognitive questionnaire with four factor analytically derived subscales (executive / speed, working memory, expressive language, episodic memory). MSCS showed a medium-sized relationship to objective cognitive impairment, and remained reliably different even when adjusting for mood. In contrast, the traditional PDQ was unrelated to objective impairment with and without adjusting for mood, despite having 2.5 times as many items as the MSCS. It may be that patients respond more thoughtfully when there are fewer items, and that MSCS items better represent the cognitive problems experienced by persons living with MS, especially given assessment of expressive language difficulty (which is missing from existing patient-report cognitive scales). The MSCS holds promise as a brief, reliable, psychometrically-robust self-report scale with good links to objective cognitive impairment.

## Data Availability

All data produced in the present study are available upon reasonable request to the authors

## Acknowledgements

The authors thank the patients, faculty, and staff of the Corinne Goldsmith Dickinson Center for Multiple Sclerosis. Special thanks to Jordyn Anderson, PsyD, Hanaan Bing-Canar, PhD, and Emily Dvorak for their work as independent raters of responses.

## APPENDIX Multiple Sclerosis Cognitive Scale (MSCS)

**Please check the box to indicate how frequently during the past month you experienced:**

|  | Never<br>(0) | Rarely<br>(1) | Some-<br>times<br>(2) | Fairly<br>Often<br>(3) | Very<br>Often<br>(4) |
| --- | --- | --- | --- | --- | --- |
| 1. losing your train of thought | ☐ | ☐ | ☐ | ☐ | ☐ |
| 2. forgetting details of a recent conversation | ☐ | ☐ | ☐ | ☐ | ☐ |
| 3. having a word “on the tip of your tongue” but with difficulty getting it out (word-finding difficulty) | ☐ | ☐ | ☐ | ☐ | ☐ |
| 4. trouble getting started, even if you had lots to do | ☐ | ☐ | ☐ | ☐ | ☐ |
| 5. forgetting what you came into the room for | ☐ | ☐ | ☐ | ☐ | ☐ |
| 6. trouble recalling what happened during the last week | ☐ | ☐ | ☐ | ☐ | ☐ |
| 7. having a sense of what you want to say, but having trouble clearly expressing your thoughts to someone else | ☐ | ☐ | ☐ | ☐ | ☐ |
| 8. taking a long time to finish things | ☐ | ☐ | ☐ | ☐ | ☐ |

## REFERENCES

1. Sumowski JF, Benedict R, Enzinger C, Filippi M, Geurts JJ, Hamalainen P, et al. Cognition in multiple sclerosis: State of the field and priorities for the future. Neurology. 2018 Feb 6;90(6):278–88.

2. Sullivan M, Edgley K, Dehoux E. A survey of multiple sclerosis: I. Perceived cognitive problems and compensatory strategy use. Canadian Journal of Rehabilitation. 1990;4:99–105.

3. Brandstadter R, Fabian M, Leavitt VM, Krieger S, Yeshokumar A, Katz Sand I, et al. Word-finding difficulty is a prevalent disease-related deficit in early multiple sclerosis. Mult Scler. 2020 Nov;26(13):1752–64.

4. Benedict RHB, Munschauer F, Linn R, Miller C, Murphy E, Foley F, et al. Screening for multiple sclerosis cognitive impairment using a self-administered 15-item questionnaire. Mult Scler. 2003 Feb;9(1):95–101.

5. Medina LD, Torres S, Alvarez E, Valdez B, Nair K V. Patient-reported outcomes in multiple sclerosis: Validation of the Quality of Life in Neurological Disorders (Neuro-QoLTM) short forms. Mult Scler J Exp Transl Clin. 2019;5(4):2055217319885986.

6. Revelle W, Revelle M. Package “psych.” The comprehensive R archive network. 2015;337(338).

7. Qin S, Nelson L, McLeod L, Eremenco S, Coons SJ. Assessing test-retest reliability of patient-reported outcome measures using intraclass correlation coefficients: recommendations for selecting and documenting the analytical formula. Qual Life Res. 2019 Apr;28(4):1029–33.

8. Langdon DW, Amato MP, Boringa J, Brochet B, Foley F, Fredrikson S, et al. Recommendations for a Brief International Cognitive Assessment for Multiple Sclerosis (BICAMS). Mult Scler. 2012 Jun;18(6):891–8.

9. Smith A. Symbol digit modalities test (SDMT) manual (revised) . Los Angeles: Western Psychological Services; 1982.

10. Brandt J, Benedict R. Hopkins Verbal Learning Test, Revised. Psychological Assessment Resources, Inc.; 2001.

11. Barnett JH, Blackwell AD, Sahakian BJ, Robbins TW. The Paired Associates Learning (PAL) Test: 30 Years of CANTAB Translational Neuroscience from Laboratory to Bedside in Dementia Research. Curr Top Behav Neurosci. 2016;28:449–74.

